# Low-concentration, low-dose lidocaine in ultrasound-guided sacral anesthesia reduces postoperative agitation and delirium in children undergoing hidden penis and hypospadias surgery: A prospective, randomized, controlled study

**DOI:** 10.1101/2025.01.30.25321386

**Authors:** Gaofeng Guo, Jiangxia Wu, Yang Zhao, Jing Zhang, Qing Yang, Jiaqiang Zhang

**Affiliations:** Department of Anesthesia and Perioperative Medicine, Henan Provincial People’s Hospital, Zhengzhou, China

## Abstract

Postoperative agitation or postoperative delirium (EA/ED) has a high incidence among pediatric patients undergoing anesthesia and surgery. In this study, we aimed to evaluate the effects of sacral anesthesia with 0.125 mL/kg of 1% lidocaine on EA/ED in children undergoing hidden penis and hypospadias surgery. Sixty children aged 1–6 years undergoing elective hidden penis or hypospadias surgery were enrolled in the study. The postoperative EA/ED score and incidence; Face, Legs, Activity, Cry, and Consolability scale; perioperative general information and vital signs; and the occurrence of adverse events were analyzed. Ultrasound-guided sacral anesthesia in children resulted in more stable vital signs, postoperative pain relief, fewer complications, and lower incidence of EA/ED. In conclusion, the use of 0.125 mL/kg of 1% lidocaine combined with laryngeal mask general anesthesia significantly reduced the incidence of EA/ED in children undergoing hidden penis and hypospadias surgery while ensuring high perioperative safety.

## INTRODUCTION

Postoperative agitation or postoperative delirium (EA/ED) is a common complication in children and is characterized by behaviors, such as crying, fear, restlessness, and disorientation occurring early in the anesthesia recovery period [1]. Studies have shown that the incidence of EA/ED among pediatric patients undergoing anesthesia and surgery is high, with mild cases accounting for approximately 35% [2, 3]. EA/ED can lead to serious consequences for children, including vomiting, aspiration, and laryngeal spasm, significantly prolonging hospital stays [4, 5]. Previous research on EA/ED in children has primarily focused on postoperative outcomes in oral and maxillofacial surgeries and otorhinolaryngological procedures [6]. Due to the rich local nerve distribution in the surgical sites of urethral hypospadias and hidden penis surgeries, along with postoperative restraints and urinary catheter placement, the incidence of postoperative EA/ED is high. However, previous studies have rarely addressed EA/ED after these surgeries. Adopting appropriate medical measures tailored to individual needs is necessary to reduce the occurrence of postoperative EA/ED in these surgeries.

Research indicates that male sex, preschool age, preoperative anxiety, sleep-disordered breathing, and postoperative pain are risk factors for EA/ED, with postoperative pain identified as an independent risk factor [3, 7, 8]. In preschool children, the sacrotuberous ligament is not ossified, enabling the safe and accurate performance of ultrasound-guided sacral anesthesia, which provides effective intraoperative and postoperative analgesia [9]. Therefore, in this study, we aimed to evaluate the effects of low concentrations and small doses of lidocaine in ultrasound-guided sacral anesthesia on EA/ED and recovery in preschool children undergoing hidden penis and hypospadias surgeries, with the goal of guiding anesthesia management for these procedures.

## MATERIALS AND METHODS

### Materials

This study was approved by the Ethics Committee of Henan Provincial People’s Hospital (No. 11,2022), and informed consent was obtained from the participants’ families. A total of 60 children undergoing their first urethral hypospadias or hidden penis repair surgery at Henan Provincial People’s Hospital between January 2023 and October 2024 were included in the study. The children were classified as American Society of Anesthesiologists grade I or II. The exclusion criteria were: (i) coagulation disorders, (ii) systemic infection or infection at the puncture site, (iii) neurological or psychiatric disorders, (iv) allergy to local anesthetics, (v) difficulty or failure in performing a sacral block, (vi) prematurity or repeated surgeries, and (vii) sacral abnormalities.

### Randomization and Blinding

Using a random envelope method, patients were divided into two groups: the ultrasound-guided sacral anesthesia combined with a laryngeal mask general anesthesia group (Group U, *n*=30) and the general anesthesia with a laryngeal mask group (Group G, *n*=30). An independent anesthesiologist, uninvolved in other aspects of the study, was responsible for recruiting patients and determining group assignments, with the group information placed in opaque envelopes. On the day of surgery, an anesthesia nurse, unaware of the purpose of the surgery, opened the envelopes to confirm group assignments and prepared the sacral block medication. A second anesthesiologist experienced in ultrasound technology performed the sacral block and managed intraoperative anesthesia, while a second anesthesia nurse recorded research data and postoperative follow-up.

## Methods

Patients underwent routine fasting before surgery. An intravenous line was established prior to entering the operating room, and standard monitoring was conducted, including electrocardiography (ECG), heart rate (HR), non-invasive blood pressure (BP), and pulse oximetry (SpO_2_). Both groups received intravenous rapid sequential induction with propofol (2–3 mg/kg), sufentanil (0.2–0.3 μg/kg), cisatracurium (0.15 mg/kg), midazolam (0.05 mg/kg), and atropine (0.01 mg/kg). Once an appropriate depth of anesthesia was achieved, a laryngeal mask of the appropriate size was inserted. The position of the laryngeal mask was confirmed by observing chest movements, end-tidal carbon dioxide (PetCO_2_) waveform changes, and auscultation of breath sounds. After confirmation, the pediatric patients were treated with mechanical ventilation. Respiratory parameters were adjusted to a tidal volume (V_T_) of 6–8 mL/kg, a respiratory rate of 15–20 breaths/min, and an inspiratory-expiratory ratio of 1:1.5, maintaining PetCO_2_ at 30–40 mmHg.

Patients in Group U were positioned on their left side in a hip-flexed, knee-flexed posture for a sacral block performed under ultrasound guidance. The puncture site was disinfected, and a sterile plastic- covered linear ultrasound probe was placed perpendicular to the posterior midline to obtain cross- sectional images. Then, scanning was conducted transversely from the coccyx toward the head (Figure 1A). Upon obtaining the transverse image of the sacral corner, the ultrasound probe was rotated 90° to capture the longitudinal image of the sacral canal. The probe was subsequently moved horizontally to the coccygeal side to observe the coccyx and then cranially to visualize S1–S5. The needle tip was observed to enter the sacral canal, ensuring no visualization of the anechoic dural sac ends. Then, the probe was adjusted centrally to display the sacrotuberous ligament and sacral hiatus. A needle was slowly advanced using a planar technique, with the needle tip visualized passing through the sacrotuberous ligament and entering the sacral canal under ultrasound imaging (Figure 1B–D). A sterile sacral puncture needle was used for the procedure, and advancement was halted as soon as the puncture needle penetrated the sacrotuberous ligament to avoid puncturing the dural sac [10]. After aspirating cerebrospinal fluid and blood, 1 mL of saline was injected. Upon observing the fluid spreading within the sacral hiatus cavity, 0.125 mL/kg of 1% lidocaine was injected at a rate of 1 mL/s. The anesthesia was considered successful if the local anesthetic was seen to diffuse within the sacral canal under ultrasound imaging; otherwise, the puncture was deemed uncertain, and the patient was excluded. Following puncture completion, a sterile protective dressing was applied to the puncture site, and the patient was placed in a supine position. Clinical operations are shown in Figure 2. In Group G, no additional treatment was performed after anesthesia induction.

**Figure 1.**
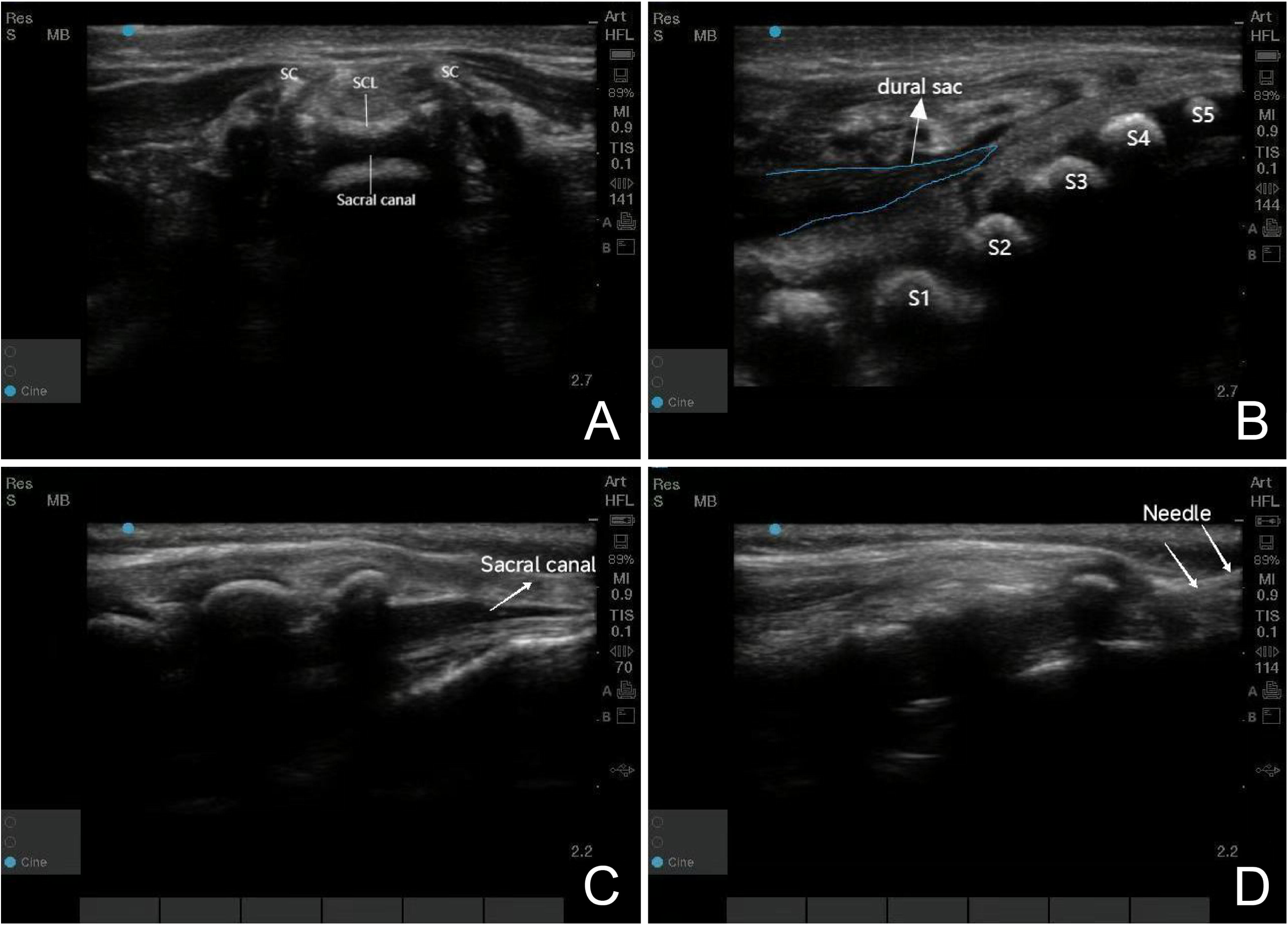
**(A)**. Cross-sectional ultrasound image of the sacral canal **(B).** Ultrasound image of the dural sac **(C).** Ultrasound image of the sacral canal in the median sagittal plane **(D).** Ultrasound image of caudal anesthesia (needle in the sacral canal)

**Figure 2.**
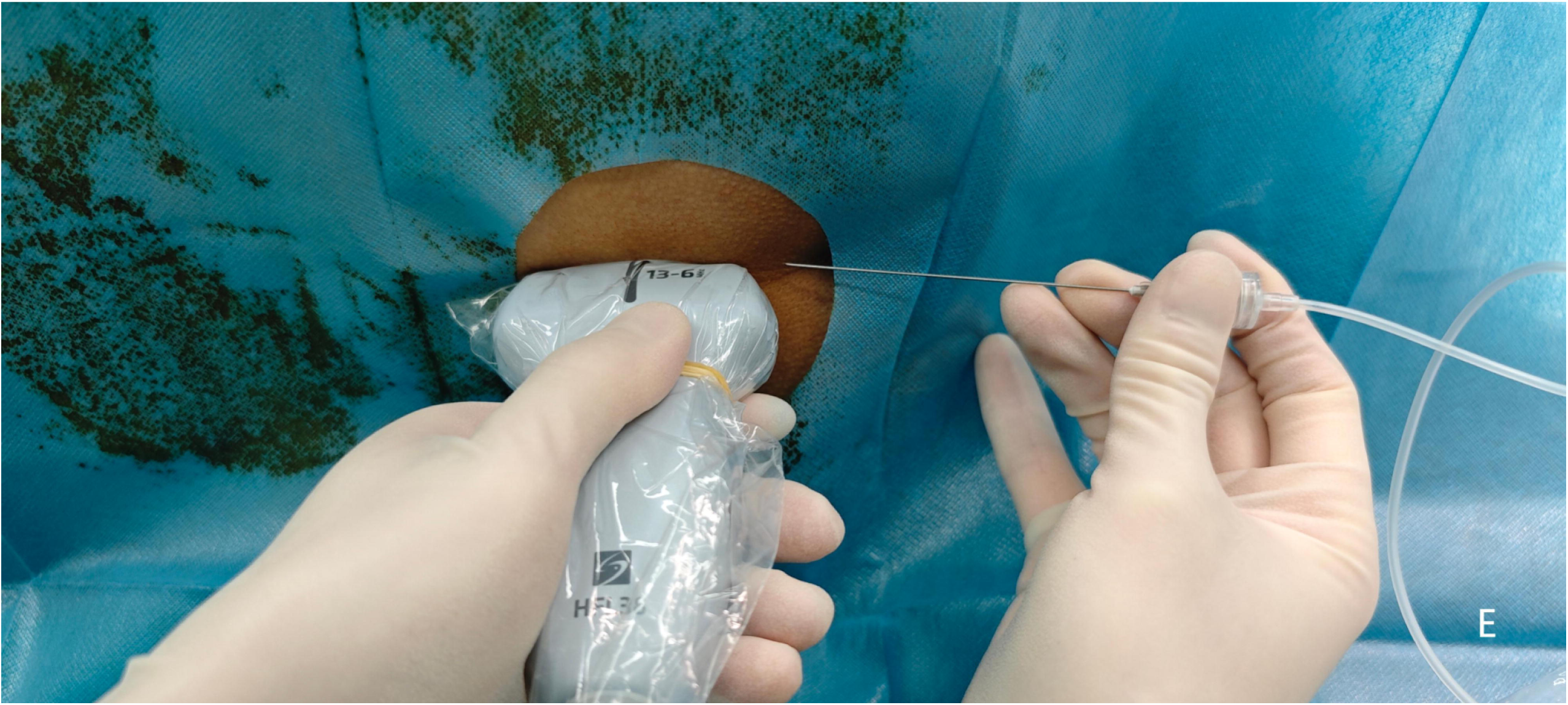
Image of the back under ultrasound-guided sacral caudal anesthesia

Intraoperatively, total intravenous anesthesia was maintained with propofol at 2–4 mg/(kg·h) and remifentanil at 0.05–0.3 µg/(kg·min), with timely dosage adjustments based on the patient’s HR and BP. The Bispectral Index (BIS) was maintained between 40–60 to ensure hemodynamic stability. If any movement occurred or significant hemodynamic fluctuations were noted (i.e., an increase in HR and mean arterial pressure [MAP] > 20% compared with pre-incision values), boluses of propofol and sufentanil were administered. The infusion of propofol and remifentanil was discontinued 5 min before the end of the surgery. The patients were transferred to the post-anesthesia care unit (PACU) after surgery.

Studies have indicated that the Richmond Agitation-Sedation Scale (RASS) and Pediatric Anesthesia Emergence Delirium (PAED) scale are effective tools for assessing EA/ED and are widely used in the PACU [11, 12]. Thus, dedicated anesthesiologists and anesthesia nurses in the PACU continued ventilatory support and vital sign monitoring using the PAED and RASS scores to evaluate postoperative agitation/delirium following extubation [13]. A PAED score >10 indicates a diagnosis of delirium and agitation [14]. Pain assessment was conducted using the Face, Legs, Activity, Cry, and Consolability (FLACC) scale for pediatric pain, which includes five components: facial expression, leg movement, activity, crying, and consolability. Each component is assigned a score of 0–2 points, with a maximum total score of 10 points; higher scores indicate greater pain severity [15].

Vital signs were monitored and recorded. The laryngeal mask was removed once the following criteria were met: the child regained consciousness, could breathe spontaneously, and could follow commands to open the mouth, lift the head, take deep breaths, or cough. Patients were discharged from the PACU once all the following criteria were satisfied: (i) the patient was awake, able to communicate, and follow commands; (ii) after removal of the laryngeal mask, the patient maintained SpO_2_ >94% for 10 min without supplemental oxygen; and (iii) pain, postoperative nausea and vomiting, and EA/ED were effectively controlled.

Sodium, potassium, magnesium, calcium, and glucose injection solutions (Jiangsu Hengrui Medicine Co., Beijing, China) were administered intraoperatively and in the PACU, according to the "4-2-1" fluid replacement principle.

If the postoperative FLACC score was ≥4 points, 15 mg/kg of acetaminophen was administered intravenously for pain relief, with a dosing frequency not exceeding every 6 h. If pain persisted after acetaminophen administration, 10 mg/kg oral ibuprofen was administered as a supplementary analgesic. If the child was comfortably asleep without disturbance during pain assessment, analgesia was considered effective [16].

### Observational Indicators

The primary observation indicator was the incidence of EA/ED. Secondary observation indicators were: surgical time; recovery time (defined as the time from the end of general anesthesia to the child’s spontaneous eye-opening); PACU observation time; HR and MAP at preoperative (T_0_), skin incision (T_1_), end of surgery (T_2_), immediately upon entering the recovery room (PACU) (T_3_), and at 5 min (T_4_), 30 min (T_5_), 60 min (T_6_), 4 h (T_7_), and 12 h (T_8_) post-extubation; FLACC, RASS, and PAED scores at T_4_, T_5_, T_6_, T_7_, and T_8_; intraoperative doses of propofol and remifentanil; and the occurrence of adverse events (bradycardia, hypoxemia, nausea, vomiting, and respiratory depression).

### Sample Size Calculation

In Group G, the reported incidence of delirium was 35% [3]. Preliminary results indicated a delirium incidence of 6%, with a power of 0.9 and a Type I error rate of <0.05, requiring 27 participants per group. Considering a 10% dropout rate, 30 participants were included in each group for observation.

Data were analyzed using SPSS version 25.0 (IBM, Armonk, NY). Normally distributed continuous variables are expressed as means ± standard deviations, with group comparisons conducted using independent samples *t*-test or repeated-measures analysis of variance. Non-normally distributed continuous variables are expressed as medians and interquartile ranges (M[IQR]), with group comparisons performed using rank-sum tests (Mann–Whitney U test). Categorical data are presented as counts (%). Group comparisons were made using *χ*^2^ tests or Fisher’s exact probability method. *P*<0.05 was considered statistically significant.

## RESULTS

Among 75 children screened for eligibility, 60 met the inclusion criteria and were randomly assigned to two groups (*n*=30): Group U and Group G. All participants completed the study, as shown in Figure 3. No significant differences were found in the patients’ general data (*P>*0.05), indicating comparability (Table 1).

**Figure 3.**
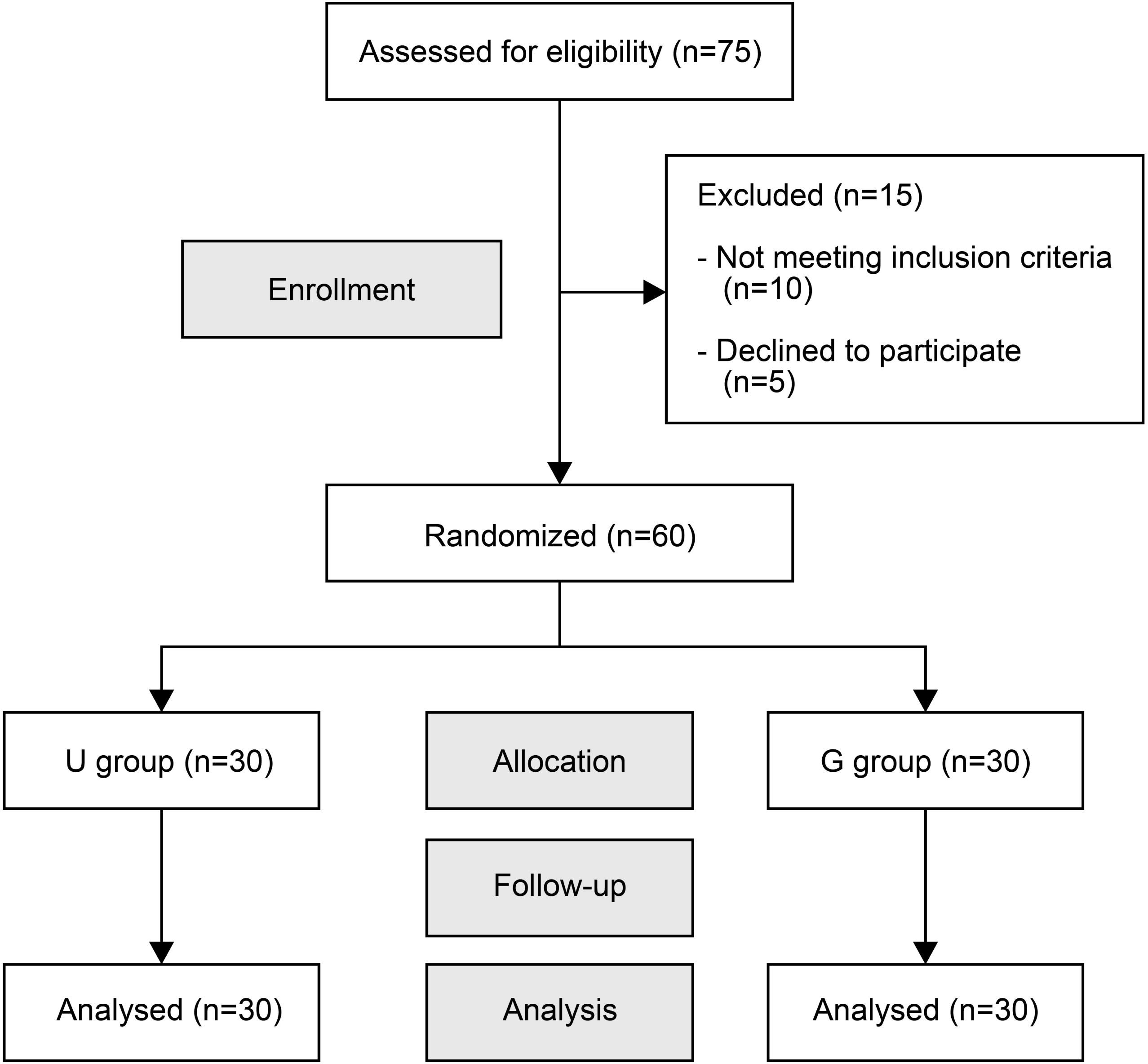
A Consolidated Standards of Reporting Trials (CONSORT) flow diagram of patient enrollment.

**Table 1.** Comparison of patient characteristics between the two groups.

| Group | Age<br>(years) | Weight<br>(kg) | Height<br>(cm) | BMI<br>(kg/m<sup>2</sup>) | Number of<br>hidden<br>penile/hypo<br>spadias | Proportio<br>n of<br>separation<br>difficultie<br>s |
| --- | --- | --- | --- | --- | --- | --- |
| U | 3.70±1.54 | 16.70±5.34 | 100.40±15.22 | 16.65±4.05 | 2/28 | 6 (20%) |
| G | 3.90±1.47 | 16.25±6.02 | 103.10±20.25 | 15.03±2.17 | 3/27 | 5 (16.7%) |
| *t/χ<sup>2</sup>* | -0.515 | 0.309 | -0.584 | 1.937 | -- | 0.111 |
| *P* | 0.608 | 0.759 | 0.562 | 0.058 | 1.000 | 0.739 |

BMI, body mass index.

The overall comparison of HR and MAP between the two groups showed significant differences over time and in interaction effects (*P*<0.05). Comparison of HR between the groups revealed significant differences (*P*<0.05), whereas the MAP comparison showed no significant difference (*P>*0.05). HR at T1, T4, as well as MAP at T4 was higher in Group G (*P*<0.05) (Tables 2 and 3)

**Table 2.**
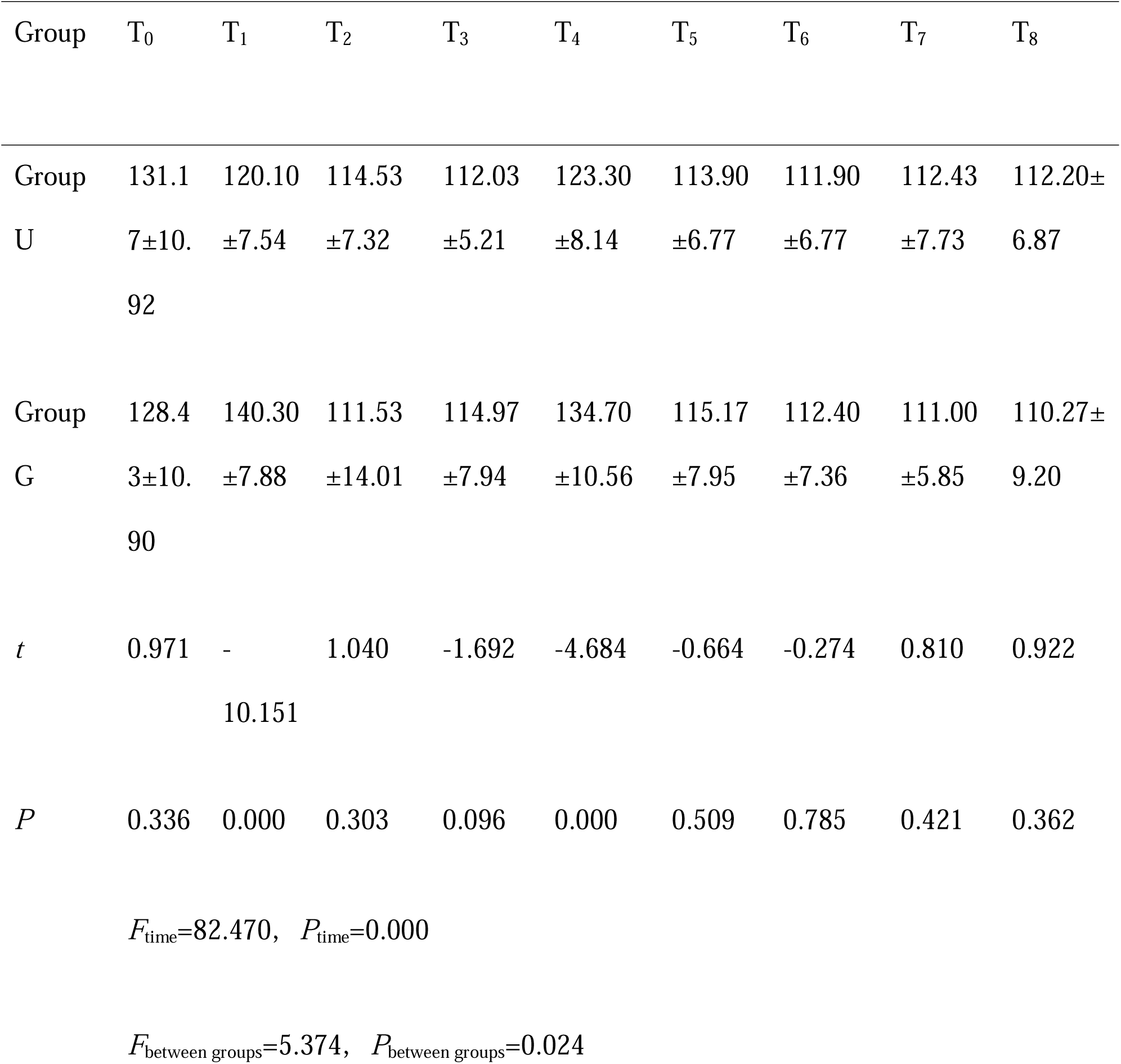
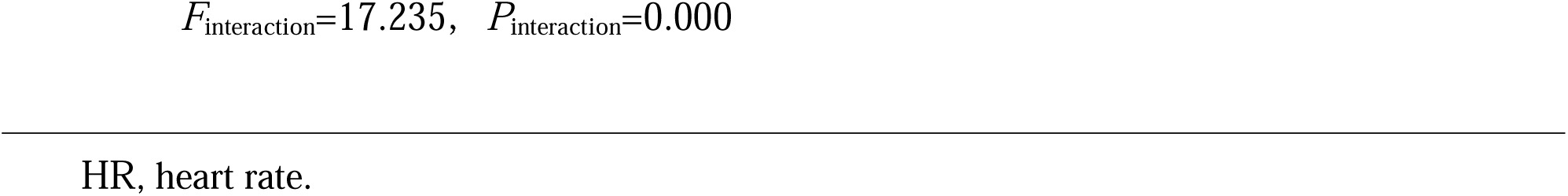
Comparison of HR at different timepoints between the two groups.

| Group | T<sub>0</sub> | T<sub>1</sub> | T<sub>2</sub> | T<sub>3</sub> | T<sub>4</sub> | T<sub>5</sub> | T<sub>6</sub> | T<sub>7</sub> | T<sub>8</sub> |
| --- | --- | --- | --- | --- | --- | --- | --- | --- | --- |
| Group | 131.1 | 120.10 | 114.53 | 112.03 | 123.30 | 113.90 | 111.90 | 112.43 | 112.20± |
| U | 7±10. | ±7.54 | ±7.32 | ±5.21 | ±8.14 | ±6.77 | ±6.77 | ±7.73 | 6.87 |
|  | 92 |  |  |  |  |  |  |  |  |
| Group | 128.4 | 140.30 | 111.53 | 114.97 | 134.70 | 115.17 | 112.40 | 111.00 | 110.27± |
| G | 3±10. | ±7.88 | ±14.01 | ±7.94 | ±10.56 | ±7.95 | ±7.36 | ±5.85 | 9.20 |
|  | 90 |  |  |  |  |  |  |  |  |
| *t* | 0.971 | - | 1.040 | -1.692 | -4.684 | -0.664 | -0.274 | 0.810 | 0.922 |
|  |  | 10.151 |  |  |  |  |  |  |  |
| *P* | 0.336 | 0.000 | 0.303 | 0.096 | 0.000 | 0.509 | 0.785 | 0.421 | 0.362 |

$F_{\text{time}}=82.470, P_{\text{time}}=0.000$
$F_{\text{between groups}}=5.374, P_{\text{between groups}}=0.024$
$F_{\text{interaction}}=17.235, P_{\text{interaction}}=0.000$
HR, heart rate.

**Table 3.**
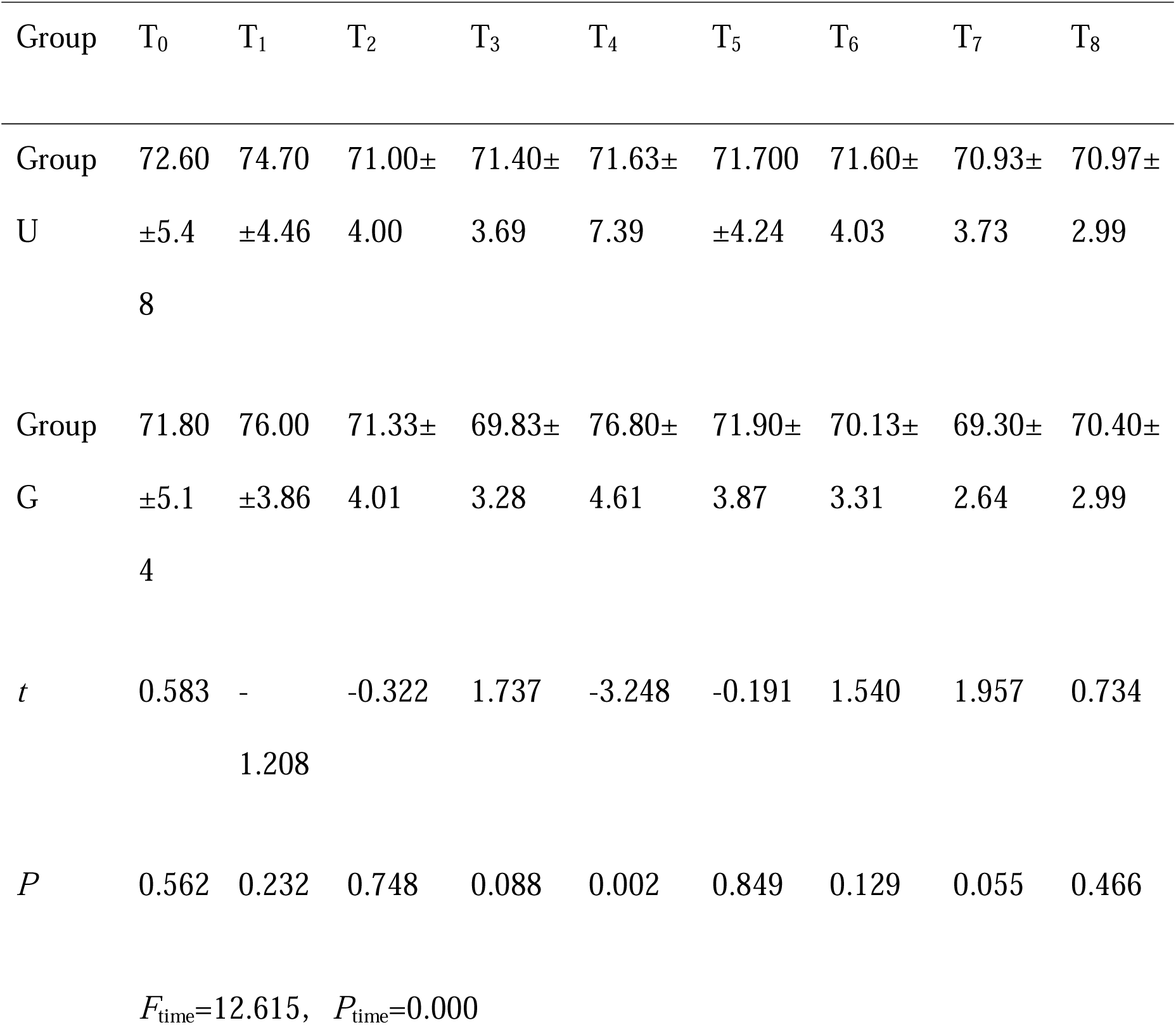
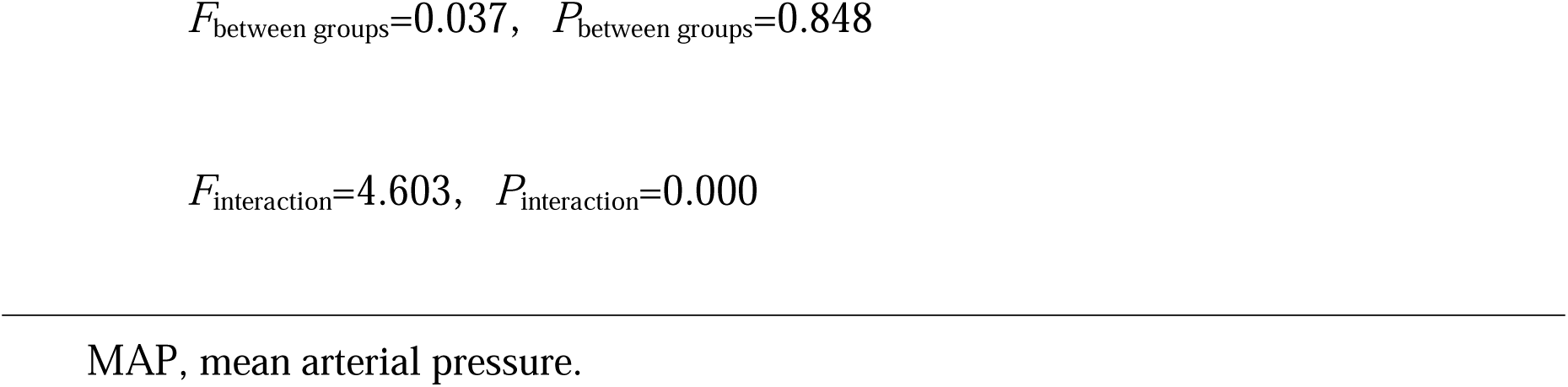
Comparison of MAP across different timepoints between the two groups.

Postoperative recovery time, PACU observation times, and intraoperative dosages of propofol and remifentanil in the two groups were higher in Group G than in Group U (*P*<0.05). No significant difference in surgical time was observed between the two groups (*P>*0.05) (Table 4).

**Table 4.** Comparison of intraoperative general information and intraoperative drug usage between the two groups.

| Group | Surgical time<br>(min) | Recovery<br>time (min) | PACU<br>observation<br>time(min) | Propofol<br>(mg) | Remifentanyl<br>(μg) |
| --- | --- | --- | --- | --- | --- |
| Group U | 96.67±27.96 | 10.93±3.40 | 47.5000<br>(27.7500) | 95.0000<br>(51.2500) | 80.0000<br>(72.5000) |
| Group G | 99.53±34.99 | 22.80±6.32 | 67.5000<br>(25.5000) | 150.0000<br>(71.2500) | 120.0000<br>(85.0000) |
| *t/Z* | -0.351 | -9.053 | -2.589 | -3.385 | -3.001 |
| *P* | 0.727 | 0.000 | 0.010 | 0.001 | 0.003 |

PACU, post-anesthesia care unit.

### Overall comparison

The FLACC, RASS, and PAED scores between the two groups showed significant differences over time and in intergroup comparisons and in interaction effects (*P*<0.05).

### Intergroup comparison

At T_4_, T_5_, and T_6_, the FLACC and PAED scores in Group U were lower than those in Group G (*P*<0.05). Similarly, at T_4_ and T_5_, the RASS scores in Group U were lower than those in Group G (*P*<0.05). At T_7_ and T_8_, the FLACC, PAED, and RASS scores, as well as the RASS score at T_6,_ showed no significant differences between the two groups (*P>*0.05), as shown in Table 5.

**Table 5.**
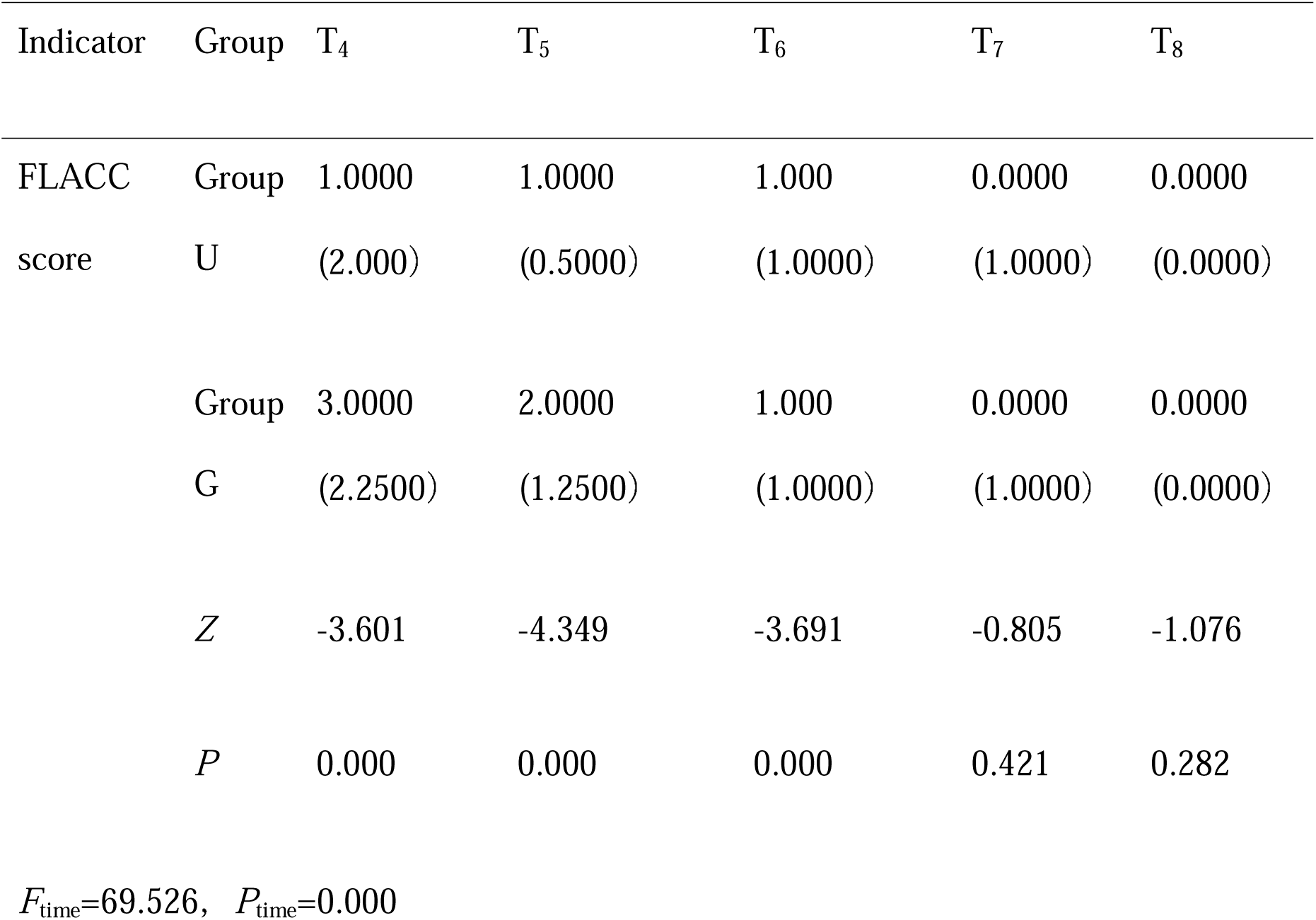
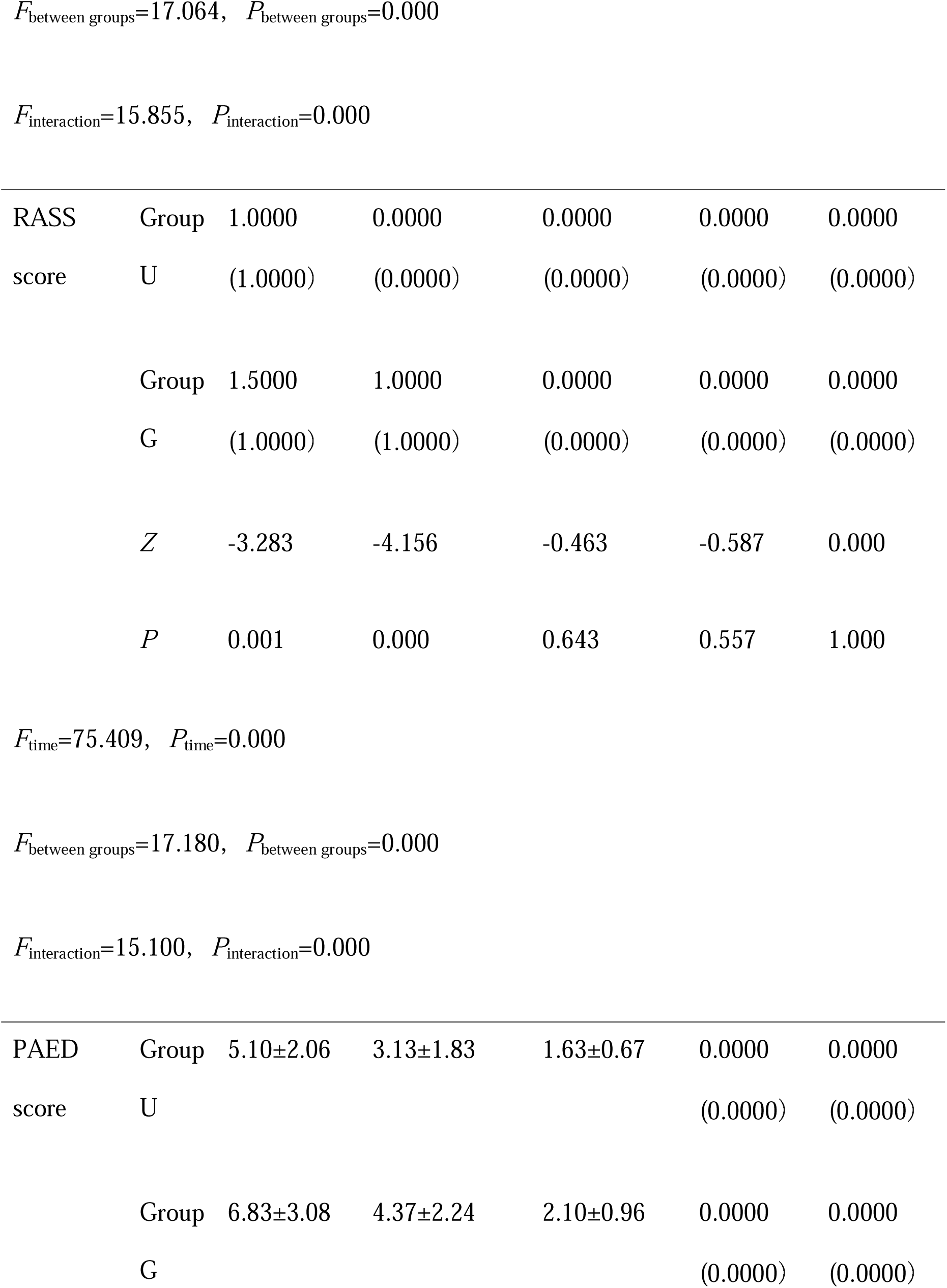
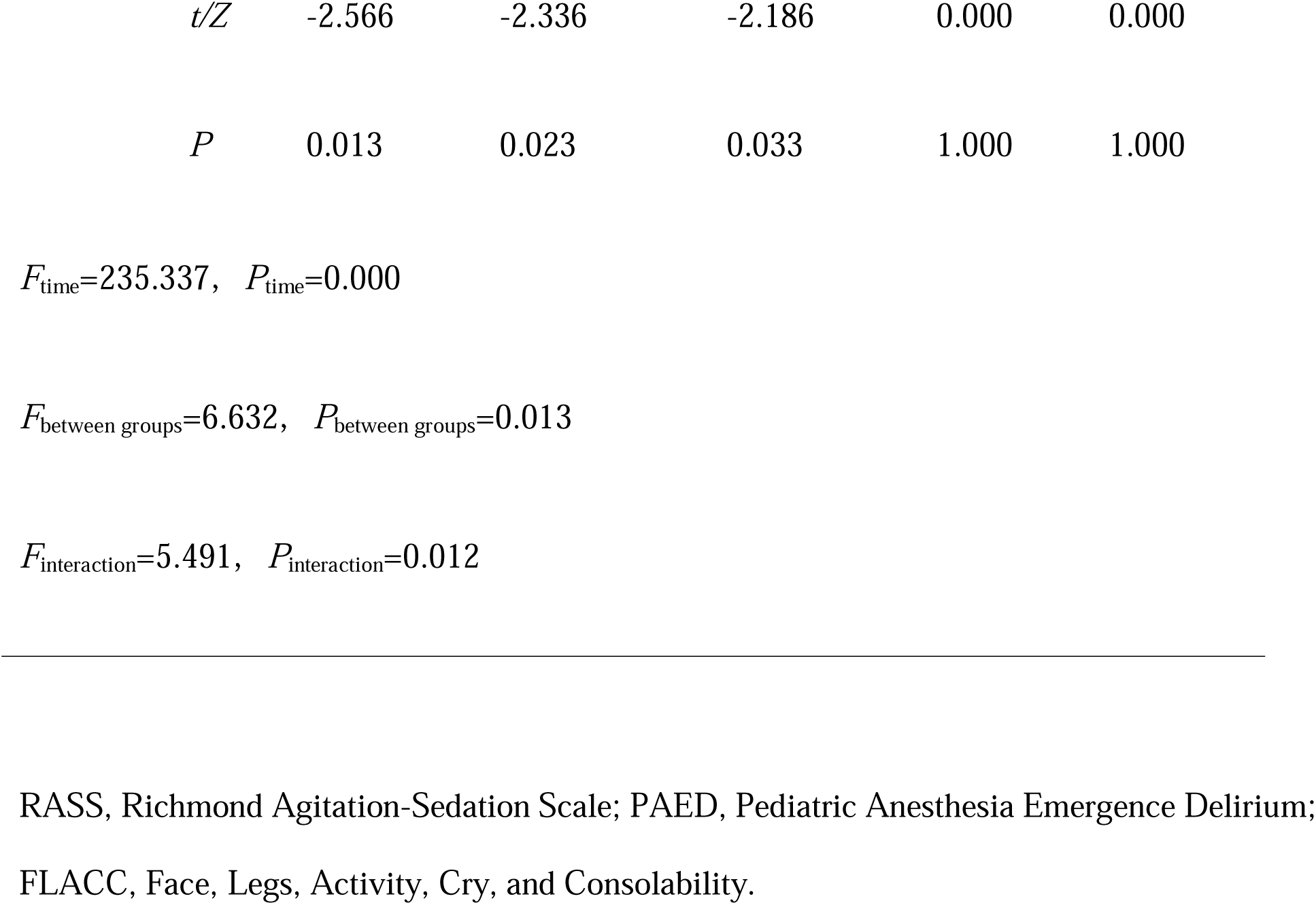
Comparison of the FLACC, RASS, and PAED scores across different timepoints between the two groups.

The incidence of respiratory depression and EA/ED during the recovery period was higher in Group G than in Group U (*P*<0.05). However, no significant differences were observed between the two groups in the incidence of bradycardia, hypoxemia, nausea, or vomiting (*P>*0.05), as shown in Table 6.

**Table 6.** Comparison of postoperative adverse events between the two groups.

| Group | Bradycardia | Hypoxemia | Nausea<br>and<br>vomiting | Respiratory<br>depression | Incidence<br>of EA/ED |
| --- | --- | --- | --- | --- | --- |
| Group U | 2 (6.7%) | 1 (3.3%) | 0 (0%) | 0 (0%) | 2 (6.7%) |
| Group G | 3 (10%) | 5 (16.7%) | 1 (3.3%) | 6 (20%) | 10<br>(33.3%) |
| $\chi^2$ | 0.000 | -- | -- | -- | 6.667 |
| *P* | 1.000 | 0.195 | 1.000 | 0.024 | 0.010 |

EA/ED, postoperative agitation and delirium.

## DISCUSSION

Emergence agitation (EA) is a postoperative complication that can occur in patients at any age but is most common in pediatric patients [17]. Emergence delirium (ED), a severe subtype of EA, is characterized by altered consciousness. The terms “EA” and “ED” are often used interchangeably [18]. Effective prevention of EA involves identifying risk factors, eliminating correctable ones, and implementing pharmacological and non-pharmacological interventions [19]. Medications commonly used to prevent EA during anesthetic recovery in children include opioids, benzodiazepines, α2 agonists, intravenous anesthetics, non-opioid analgesics, short-acting midazolam, and dexmedetomidine [20, 21]. However, these medications may prolong post-anesthesia sedation, delaying awakening and causing other adverse reactions [22].

Smooth anesthesia recovery and effective perioperative analgesia can reduce the incidence of EA/ED [5]. Sacral anesthesia is a widely used analgesic technique in pediatric patients. However, anatomical variations in the sacrum can result in failure rates of up to 30% when surface landmark-based puncture techniques are used [23]. Ultrasound guidance improves accuracy by visually confirming drug diffusion. A concentration of 0.25%–0.5% bupivacaine at 1 mL/kg or 0.1%–0.375% ropivacaine at 1 mL/kg is commonly used for pediatric sacral anesthesia [16, 24, 25]. However, these local anesthetics may increase the incidence of postoperative motor block, lower limb numbness, and urinary retention, thereby extending the child’s recovery time. Moreover, the small sacral space in children and the loose attachment of epidural fat and fascia to the nerves enhance the diffusion of local anesthetics, potentially resulting in excessively high block levels [9, 26]. Recent studies have suggested that the minimum effective concentration for an ultrasound-guided femoral nerve block is 0.93% [27]. Other studies recommend 5 mL of 1% lidocaine for sacral anesthesia [28, 29], while 0.125 mL/kg of local anesthetic provides a block range from L5 to S1 [30]. Therefore, in this study, we aimed to investigate the effects of 0.125 mL/kg of 1% lidocaine for sacral anesthesia on postoperative EA/AD and recovery in children undergoing hidden penises and hypospadias surgery.

In this study, ultrasound-guided sacral anesthesia reduced the incidence of EA/ED, with significant differences in FLACC and PAED scores at T_4_, T_5_, and T_6_, which were lower in Group U. Similarly, the RASS scores were lower in Group U at T_4_ and T_5_. These findings suggest that 0.125 mL/kg of 1% lidocaine used in ultrasound-guided sacral anesthesia effectively reduces the incidence of EA/ED in preschool children undergoing hypospadias and hidden penis surgeries. The use of a low concentration and small dosage of lidocaine in ultrasound-guided sacral anesthesia significantly decreased the incidence of EA/ED due to the following reasons. First, ultrasound guidance ensures accurate localization and real-time monitoring of local anesthetic diffusion, facilitating complete drug dispersion, while also excluding patients with anatomical variations that could compromise the success and efficacy of sacral anesthesia. Second, the nerves innervating the area of interest are within the effective range of sacral anesthesia dosages used in this study [30]. Third, the surgery duration was relatively short, and the analgesic effect of sacral anesthesia reportedly lasts up to 18 h [16], providing excellent postoperative analgesia. Fourth, upon transfer to the PACU, patients remained under anesthesia, allowing the sacral effect to reduce noxious stimuli, facilitating a smoother transition for patients [5]. Finally, the simultaneous recovery of brain function enhanced patients’ self-control abilities, which in turn reduces the occurrence of EA/ED [31, 32]. Additionally, the results revealed no significant difference in EA/ED scores at 60 min post-extubation. EA/ED are self-limiting symptoms that typically appear within the first 30 min postoperatively and significantly decrease afterward [3]. In this study, however, a reduction in scores was observed only after 60 min, likely because PACU staff administered appropriate medications to relieve symptoms, thereby extending the observation and assessment period. Although the self-limiting nature of EA/ED raises questions regarding proactive interventions, patients with EA/ED face increased risks of catheter removal, wound infection, and falls during the recovery period, placing a significant strain on PACU anesthesia providers and nursing staff. Furthermore, children experiencing EA/ED may exhibit behavioral and habitual changes lasting up to 4 weeks postoperatively [33]. Therefore, prompt and effective interventions are necessary. Consistent with previous studies [21, 34], the findings of this study show that pain relief in children corresponded with a reduction in EA/ED scores.

Preschool children have small sacral volumes. Koo et al. evaluated the diffusion of ropivacaine using fluoroscopy, demonstrating that 0.5, 1.0, and 1.25 mL/kg of local anesthetic diffused to the levels of L2, T12, and T10, respectively [35]. Other studies suggest that sacral injection of local anesthetics poses a risk of inducing intracranial hypertension [36], especially as the degree of increased intracranial pressure appears to be related to the volume of the local anesthetic used [37]. Therefore, using a minimal effective volume of local anesthetic alleviates concerns regarding elevated intracranial pressure. Additionally, unnecessary increases in anesthetic levels are strongly associated with hypotension and bradycardia. In the current study, the dosages and concentrations used did not cause symptoms of increased intracranial pressure, hypotension, or bradycardia associated with excessively high block levels. Additionally, studies have found that higher concentrations of lidocaine are significantly neurotoxic, while lower concentrations and doses are associated with reduced toxicity [38]. Therefore, appropriate low-concentration and low-dose anesthetic agents are essential for sacral applications. The lidocaine used in this study was within a safe and effective dosage and concentration range, without excessive dosages or high concentrations that could lead to adverse reactions, thereby ensuring safety.

In this study, HR was higher in Group G at T_1_ and T_4_, and MAP was higher in Group G at T_4_, than those in Group U. These findings may be attributed to ultrasound-guided sacral anesthesia providing effective perioperative analgesia, minimizing adverse stimuli during skin incision and laryngeal mask removal, and maintaining more stable hemodynamics.

In this study, the dosages of propofol and remifentanil used in Group U were lower than those used in Group G. We hypothesize that the sacral block may provide effective analgesia comparable to that of opioids. Achieving an appropriate depth of anesthesia allowed for reduced intraoperative anesthesia use. Despite greater opioid use in Group G, the incidence of nausea and vomiting showed no significant differences between the two groups. This may be attributed to the surgery duration, which limited opioid retention, combined with the rapid metabolism of remifentanil, reducing the likelihood of accumulation. Additionally, nausea, defined as a sensation accompanied by a tendency to vomit, is challenging for preschool children to express [39], potentially leading to underestimation. The lower incidence of postoperative respiratory depression in Group U may be attributed to the higher FLACC scores in Group G, which necessitated supplemental analgesia that likely suppresses respiration during recovery. While other adverse events did not show statistical significance, Group U experienced better response to analgesia, suggesting that the analgesia technique to be safer and more beneficial for children.

This study has some limitations. First, this was a single-center study with a small sample size. Second, the long-term impact of EA/ED on children’s behavioral abnormalities and their relationship requires further longitudinal studies. Third, the optimal effective concentration of 1% lidocaine in sacral anesthesia is yet to be determined. Finally, the efficacy of other local anesthetics for sacral applications was not evaluated.

In summary, 0.125 mL/kg of 1% lidocaine in sacral anesthesia combined with laryngeal mask general anesthesia significantly reduces the incidence of EA/ED in children undergoing hidden testicular and hypospadias surgeries while ensuring high perioperative safety.

## Data Availability

The raw data supporting the conclusions of this article will be made available by the authors on request

## Acknowledgments

We would also like to thank Editage (www.editage.cn) for the English language editing.

